# Assessment of Dosimetric Accuracy and Plan Quality in Prostate SBRT: Retrospective Evaluation of 36.25 Gy in 5 Fractions Using RapidArc on Varian Millennium MLC with MLC Parameter Analysis

**DOI:** 10.1101/2025.07.16.25331530

**Authors:** M Ajithkumar, Subhankar Show, Aaditya Prakash

## Abstract

**Introduction:** Prostate cancer is one of the most commonly diagnosed cancers among men worldwide. Advances in radiotherapy, particularly Stereotactic Body Radiotherapy (SBRT), have enabled ultra-hypofractionated treatment schedules that enhance tumor control while reducing treatment time. This study focuses on evaluating the dosimetric accuracy and plan quality of prostate SBRT using RapidArc technology on a Varian Millennium Multileaf Collimator (MLC) system.

**Materials and Methods:** A total of 24 patients with localized prostate adenocarcinoma received SBRT with a prescribed dose of 36.25 Gy in five fractions. Treatment planning was performed using Eclipse v15.6 with Acuros XB algorithm, utilizing three 6 MV flattening filter-free (FFF) arcs. Planning Target Volume (PTV) coverage, OAR doses, Paddick Conformity Index (PCI), Gradient Index (GI), and Monitor Units (MU) were analyzed. Multileaf collimator motion was evaluated through log files, including leaf speed and position dynamics. Quality assurance was performed using electronic portal imaging devices (EPID) and gamma pass rate evaluation.

**Results:** The mean PTV D_95_ was 35.80 ± 0.46 Gy, with mean Dmax and Dmean being 39.80 ± 1.05 Gy and 37.10 ± 0.40 Gy, respectively. V_95%_ averaged 99.13 ± 1.14%, confirming adequate coverage. Slight violations of rectum and bladder Dmax constraints were observed but remained clinically acceptable. The mean PCI and GI were 0.905 ± 0.18 and 3.08 ± 0.16, respectively. Gamma pass rates exceeded 99.6% for 2%/2 mm criteria. MLC leaf speed remained below the 2.5 cm/s threshold, ensuring mechanical safety and dose delivery accuracy.

**Conclusion:** Prostate SBRT delivered via RapidArc on a Varian Millennium MLC system demonstrated high plan conformity, efficient delivery, and acceptable OAR sparing. The integration of intra-fraction CBCT improved setup accuracy, while MLC analysis confirmed mechanical precision. This approach supports the clinical adoption of RapidArc-based SBRT as a feasible and effective option for localized prostate cancer treatment.

## 1. Introduction

Prostate cancer remains an important global health challenge, particularly affecting aging male populations worldwide. Advances in radiotherapy have revolutionized its management, with technological innovations enabling precise targeting and dose escalation to improve tumor control while minimizing adverse effects. Among these developments, SBRT has emerged as a promising treatment modality, delivering high biological doses in five fractions, thus offering a more convenient and resource-efficient alternative to conventional radiotherapy regimens[1-2].

The radiobiological rationale for hypofractionation in prostate cancer is grounded in its low alpha/beta ratio (∼1.5 Gy) [1-14], which suggests that larger doses per fraction can enhance tumor control and potentially spare normal tissues. Supporting this approach, clinical trials such as PACE-B and HYPO-RT-PC have demonstrated favorable outcomes with regimens like 36.25 Gy in five fractions, showing good tolerability and promising efficacy [3]. These findings align with the trend toward ultra hypofractionation, which aims to optimize treatment efficacy while reducing treatment duration, healthcare costs, and patient burden.

However, the success of such high-dose, few-fraction treatments hinges critically on meticulous dosimetric planning. Precise delineation of target volumes and organs at risk (OARs)—notably the bladder, rectum, and penile bulb—is essential to maximize tumor coverage and minimize radiation exposure to surrounding normal tissues. Techniques like advanced image-guided planning used to achieving these goals [1-3].

While prior studies have explored SBRT for prostate cancer using RapidArc, this work uniquely combines conventional dosimetric evaluation with a detailed analysis of multileaf collimator (MLC) leaf speed dynamics, an understudied aspect of treatment delivery stability. By characterizing MLC motion through leaf speed distributions, we provide novel insights into the mechanical reliability of plan delivery. Additionally, this study evaluates the impact of daily intra-fraction CBCT-based image guidance on ensuring precise dose delivery in a high-dose, ultra-hypofractionated regimen (36.25 Gy in five fractions). This integrated dosimetric and mechanical approach offers a comprehensive assessment of treatment quality, paving the way for refined clinical protocols and enhanced safety in prostate SBRT.

Given the ongoing evolution of prostate radiotherapy protocols and the increasing adoption of SBRT, our study aims to evaluate the plan quality and adherence to dose constraints in delivering prostate SBRT in five fractions. This research will contribute to establishing standardized dosimetric guidelines, ensuring safe and effective treatment while preserving the quality of life for patients. As the landscape of prostate cancer treatment continues to advance, precise and validated dosimetric planning remains the cornerstone for translating promising clinical outcomes into routine practice.

## 2. Materials and Methods

### 2.1 Study Design and Patient Selection

This retrospective cohort study included a total of 24 patients diagnosed with localized prostate cancer between [2022–2025] .The sample size was based on all eligible non-metastatic cases treated during the study period. Diagnosis was established using serum prostate-specific antigen (PSA) testing, digital rectal examination (DRE), transrectal ultrasound-guided biopsy, and computed tomography (CT) imaging to confirm localized disease. All patients received SBRT in exactly five fractions, following established ultra-hypofractionation protocols for prostate cancer.

#### Inclusion Criteria

1. Male patients aged 18 years or older,
2. Histologically confirmed prostate adenocarcinoma,
3. Clinical staging indicating localized disease (e.g., T1c-T3a),
4. No evidence of metastatic disease,
5. Prostate volume within the range suitable for SBRT (e.g., 90-120 cc),
6. Adequate organ function and performance status suitable for radiotherapy.

#### Exclusion Criteria

1. Prior pelvic radiotherapy,
2. Significant comorbidities contraindicating SBRT,
3. Presence of metastatic or locally advanced disease beyond T3a,
4. Inability to tolerate immobilization or imaging procedures necessary for SBRT.

### 2.2 Imaging and Target Volume Delineation

All patients underwent CT simulation with a 1 mm slice thickness using a linear accelerator. Immobilization was performed in the supine position with knee and foot supports. Patients followed bladder filling and rectal emptying protocols to reduce internal organ motion. CT images were imported into the Eclipse 15.6 treatment planning system (Varian Medical Systems, Palo Alto, CA, USA) [1].

The gross tumor volume (GTV) was delineated based on MRI and PET/CT imaging, particularly utilizing Ga-68 PSMA PET scans to identify dominant intraprostatic lesions with high tracer uptake. The clinical target volume (CTV) included the prostate and seminal vesicles, with a 3 mm isotropic expansion to account for potential microscopic extension. A planning target volume (PTV) was then generated by adding an additional 3 mm isotropic margin around the CTV to compensate for setup uncertainties and ensure accurate dose delivery.

Organs at risk (OARs), such as the bladder, rectum, penile bulb, femoral heads, and small bowel, were contoured following standard guidelines. The rectum extended from the ischial tuberosities to the rectosigmoid flexure; the bladder encompassed the entire organ. Additional OARs, including the penile bulb and femoral heads, were contoured for dose monitoring [15-17].

### 2.3 Treatment Planning and Delivery

Treatment plans were developed in Eclipse 15.6 (Varian Medical Systems, Palo Alto, CA, USA) using the Acuros XB(AXB) algorithm for dose calculation [10, 15-20]. The prescribed dose was 36.25 Gy delivered in five fractions. Plans aimed for at least 95% of the PTV volume to receive the prescribed dose (V_95_ ≥ 95%), with the isodose line set at 95% to cover the target, allowing hotspots up to approximately 111% within the PTV.

The optimization utilized three full arcs with 6 MV flattening filter-free (FFF) photon beams configured as Volumetric Modulated Arc Therapy (VMAT). The plan prioritized conformality and OAR sparing, with the dose distribution normalized to keep the 95% isodose line encompassing the PTV. Hotspots within the target were permitted to reach up to 111% of the prescribed dose, provided they did not exceed OAR constraints. Dose constraints included maximum doses of 40 Gy to the penile bulb and 38 Gy to the rectum and bladder.

DVH analyses evaluated key metrics: D_95_, Dmax, Dmean, and V_95_ for the target. The primary target coverage metric was V_95_, representing the percentage volume of the PTV receiving at least 95% of the prescription dose.

### 2.4 Dosimetric Parameters and Plan Quality Metrics

The Paddick Conformity Index (PCI), a widely used measure of plan conformity [5, 9-12], was calculated as follows:

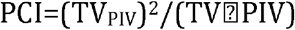

Where:

TV_PIV_: Target volume covered by prescription isodose volume

TV: Target volume

PIV: Prescription isodose volume

A PCI value close to 1 indicates optimal conformity between the prescribed dose distribution and the target volume.

#### Gradient Index (GI)

Defined as:

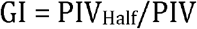

where PIV_half_ represents the prescription isodose volume at half the prescription isodose (e.g., at 40 %), and PIV, the full prescription isodose volume (e.g., at 80 %). This aligns with the standard used in ICRU Report 91 for assessing dose fall-off outside the target. A lower GI (Typically 3-4) reflects a steeper dose gradient, indicating better normal tissue sparing.

### 2.5 MLC Parameters

MLC model used was the Varian Millennium 120, featuring a 5 mm leaf width at isocenter in the central 20 cm of the field, optimized for precise beam shaping. Maximum leaf speed was verified to remain within machine specifications (e.g., up to 2.5 cm/s) during dynamic delivery [5]. MLC position data were recorded at 20 ms intervals during treatment delivery. These high-frequency log files enabled detailed analysis of leaf motion patterns, positional stability, and dynamic adaptation throughout the arc. The mean leaf speed was computed from frame-to-frame positional shifts to ensure accurate and consistent dose delivery (see Figures 6 and 7).

### 2.6 Treatment Delivery Data

#### MU Count

Total monitor units delivered per plan.

#### MLC Motion Patterns

Assessed from logged positional data to evaluate plan complexity(Total MU/cGy) and delivery efficiency.

#### Treatment Times

Total beam-on time recorded to evaluate treatment efficiency

### 2.7 Quality Assurance and Verification

The advanced irradiation techniques in a radiotherapy clinic require extensive dose verification measures that go beyond current routine clinical practice. Amorphous silicon electronic portal imaging devices (a-Si EPIDs) (e.g., Varian aS1200, Varian Medical Systems, Palo Alto, CA, USA) [15]. Pre-treatment verification was performed using the a-Si EPID(Varian aS1200, Varian Medical Systems, Palo Alto, CA, USA) system, with gamma passing rates evaluated at 3%/2 mm and 2%/2 mm criteria to ensure plan accuracy. The Varian linear accelerator automatically logs thirteen standard delivery parameters via trajectory files, including monitor units (MU), gantry and collimator angles, MLC positions, jaw settings, dose rate, and couch position. Among these, MU, MLC motion, and gantry angle were selected for analysis due to their direct impact on delivery accuracy. Treatment delivery times were optimized to improve efficiency without compromising plan quality.

### 2.8 Statistical Analysis

All dosimetric parameters, including D_95_, Dmax, Dmean, V_95_, and OAR doses, were statistically analyzed. Descriptive statistics summarized plan quality, and comparisons were made to assess consistency across patients. A significance threshold of p<0.05 was applied.

Treatment plans were developed in Eclipse 15.6(Varian Medical Systems, Palo Alto, CA, USA). The plans used three 6 MV FFF arcs, with the isodose line set at 95% to ensure adequate target coverage, allowing hotspots up to 111% [3,5,7,9-13]. The approach emphasizes robust tumor coverage (V_95_ ≥ 95%) while maintaining strict OAR constraints, aligning with current clinical practices for safe, effective ultra hypofractionated prostate radiotherapy.

## 3. Results

### Patient Demographics

A total of 24 patients were included in the study. The demographic characteristics are summarized in Table 1. The mean age was 50.0 ± 6.7 years, with an age range from 30 to 70 years. All patients were male and presented with localized disease at diagnosis.

**TABLE 1:** Patient Demographics.

| Characteristic | Number / Details |
| --- | --- |
| Total Patients | 24 Patients |
| Age (years), mean ± SD | 50.0 ± 6.7 |
| Age Range | 30 – 70 |
| Gender | Male (24 Patients) |
| Disease Staging (clinical T stage) |  |
| ☐ T1c | 6 patients (25%) |
| ☐ T2a | 8 patients (33%) |
| ☐ T2b | 5 patients (21%) |
| ☐ T3a | 5 patients (21%) |
| Prostate Volume Range | 90 – 120 cc |

Summary of baseline demographic and clinical characteristics for 24 male patients with localized prostate cancer treated with stereotactic body radiotherapy (SBRT) to a total dose of 36.25 Gy in 5 fractions. Clinical T-stage distribution includes T1c to T3a disease.

Table 2 demonstrates the Statistical Summary of Dosimetry Metrics

**TABLE 2:** Statistical Summary of Dosimetry Metrics.

| Structure | Metric | Average ± SD | Unit | Constraint |
| --- | --- | --- | --- | --- |
| PTV | Dmean | 37.10 ± 0.40 | Gy | - |
| PTV | Dmax | 39.80 ± 1.05 | Gy | - |
| PTV | D<sub>95</sub> | 35.80 ± 0.46 | Gy | ≥ Prescription (36.25 Gy) |
| PTV | Dmin | 32.51 ± 0.52 | Gy | - |
| PTV | V<sub>95</sub> (%) | 99.13 ± 1.14 | % | ≥ 95% |
| Bowel Bag | Dmax | 21.55 ± 1.12 | Gy | < 35 Gy |
| Bowel Bag | V<sub>20Gy</sub> | 25.50 ± 69.45 | cc | < 50 cc |
| Bladder | Dmax | 38.76 ± 0.86 | Gy | < 38 Gy |
| Bladder | V<sub>18Gy</sub> | 40.91 ± 13.74 | % | < 50% |
| Rectum | Dmax | 38.48 ± 0.70 | Gy | < 38 Gy |
| Rectum | V<sub>18Gy</sub> | 44.48 ± 9.50 | % | < 50% |
| Femoral Head (Rt) | V<sub>30Gy</sub> | 0.00 ± 0.00 | cc | < 5 cc |
| Femoral Head (Lt) | V<sub>30Gy</sub> | 0.00 ± 0.00 | cc | < 5 cc |
| Penile Bulb | Dmax | 38.72 ± 0.81 | Gy | < 40 Gy |
| Penile Bulb | V<sub>30Gy</sub> | 47.30 ± 6.69 | % | < 50% |

Dmean (Mean Dose): The average dose received by the entire PTV is approximately 37.10 Gy, with a small variation (±0.40 Gy). This indicates that, on average, the target volume receives slightly above the prescribed dose (36.25 Gy), which can be intentional to ensure coverage. Dmax (Maximum Dose): The highest dose within the PTV is about 39.80 Gy (±1.05 Gy), reflecting dose hotspots or inhomogeneities, which are common in SBRT to boost tumor control. D_95_: The dose received by 95% of the PTV is approximately 35.80 Gy (±0.46 Gy). It is slightly below the prescribed dose (36.25 Gy), but still generally acceptable for ensuring adequate coverage, as it ensures most of the target gets the intended dose. Dmin (Minimum Dose): The lowest dose within the PTV is around 32.51 Gy, indicating some dose heterogeneity but unlikely to compromise treatment efficacy.

Bowel Bag V_20Gy_: A mean volume of 25.50 ± 69.45 cc received 20 Gy, with a notably high standard deviation. This variability is likely due to inter-patient anatomical differences and fluctuations in bladder filling, which can affect bowel positioning. These findings underscore the importance of standardized bowel and bladder preparation protocols to minimize dose to the bowel and improve plan consistency. Bladder/Rectum Dmax violate constraints (p<0.05) [14, 17-19], Bowel Bag and Penile Bulb are within limits (p>0.05) [16]. Where slight exceedances of predefined dose constraints are observed for organs such as the bladder and rectum. Specifically, a slight exceedance indicates that the dose to certain organs is marginally above the established safety limits, but these levels remain close to the constraints (see Table 2).In practice, small violations may be acceptable if they do not significantly increase the risk of toxicity or adverse effects. This approach allows for flexibility in treatment planning, enabling clinicians to prioritize effective tumor coverage and overall patient safety while acknowledging minor deviations from ideal constraints.

Figure 1 illustrates the dose-volume histogram for a representative patient, demonstrating adequate target coverage and organ sparing. Figures 2, 3, and 4 show views of the 50% dose distribution, demonstrating conformal coverage of the prostate (outlined) with rapid dose fall-off outside the target.

**FIGURE 1:**
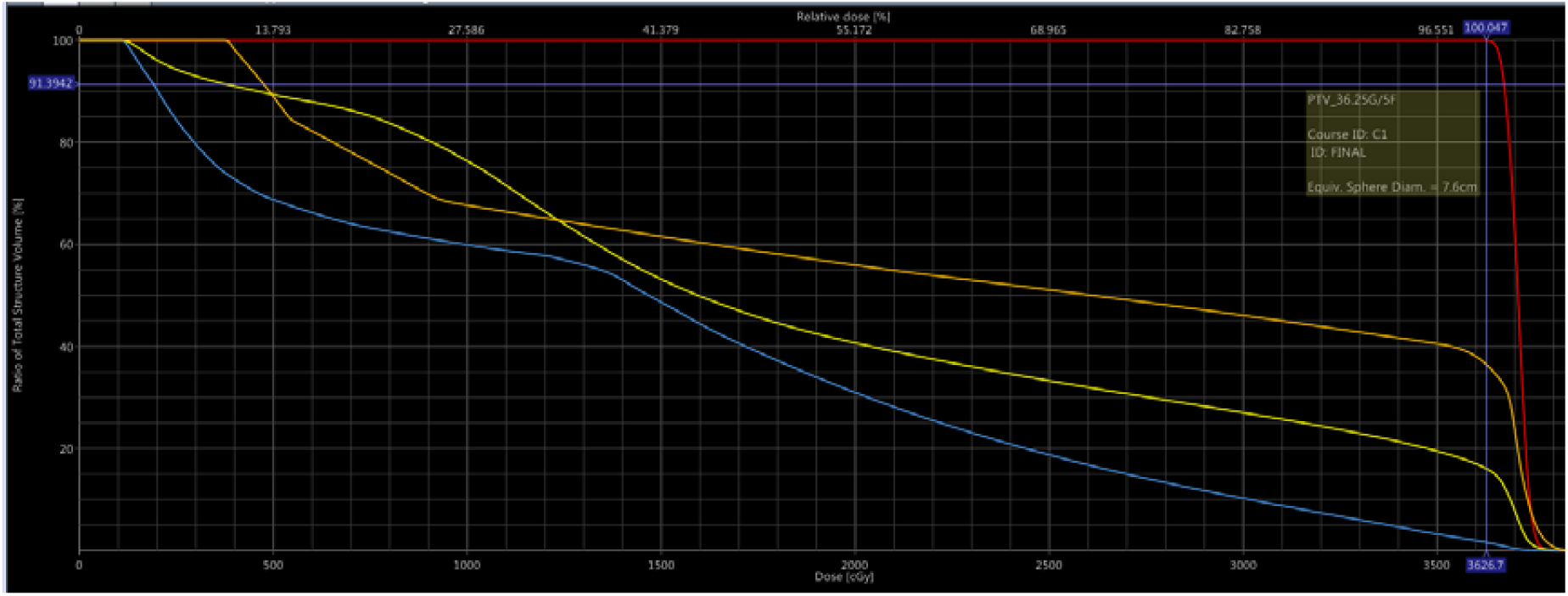
Dose-volume histogram for a representative patient demonstrating target coverage (PTV) and sparing of OARs (bladder, rectum, penile bulb). The dashed line indicates the prescription dose of 36.25 Gy.

**FIGURE 2:**
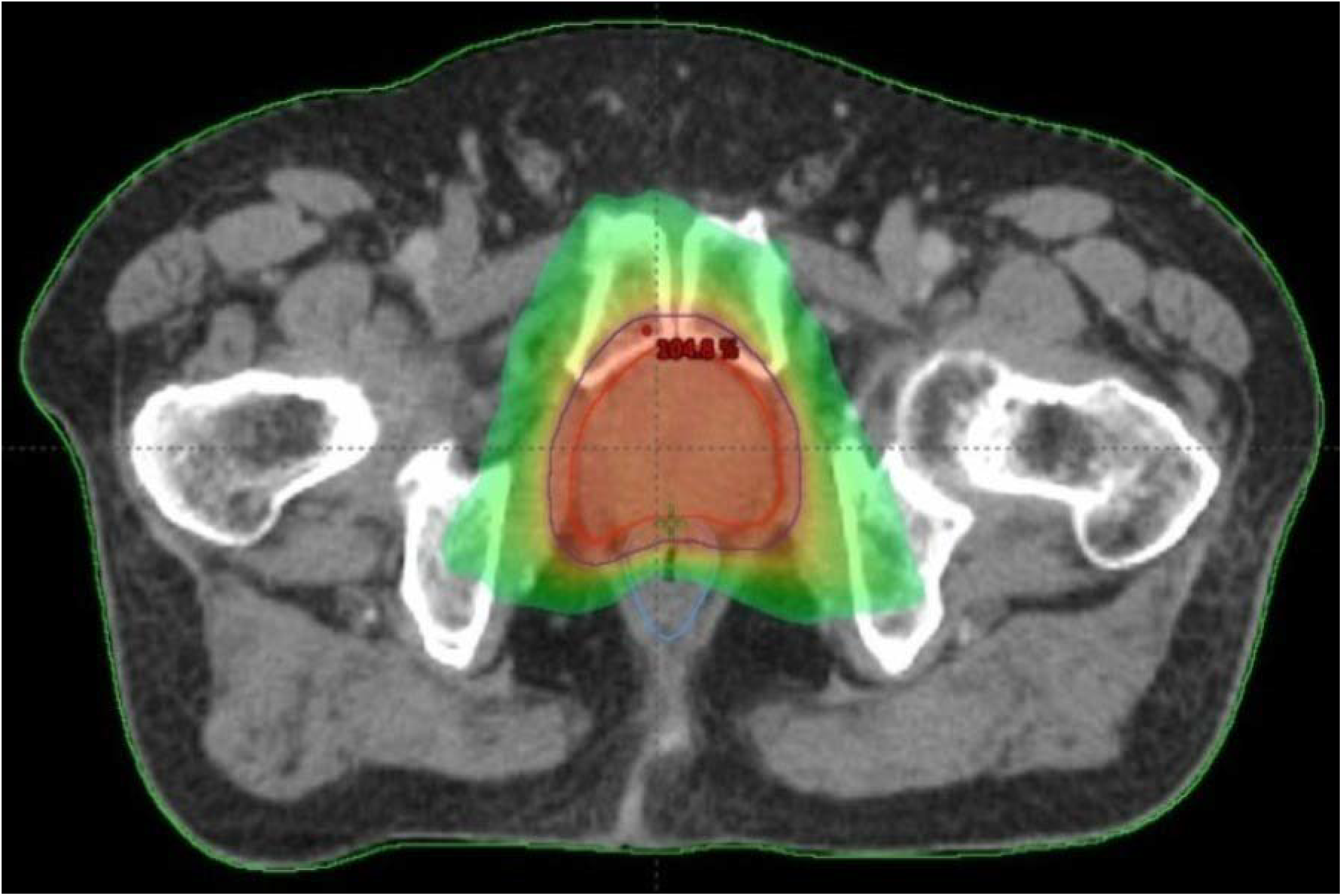
Pre-treatment axial CT image of the prostate showing 50% isodose line distribution.

**FIGURE 3:**
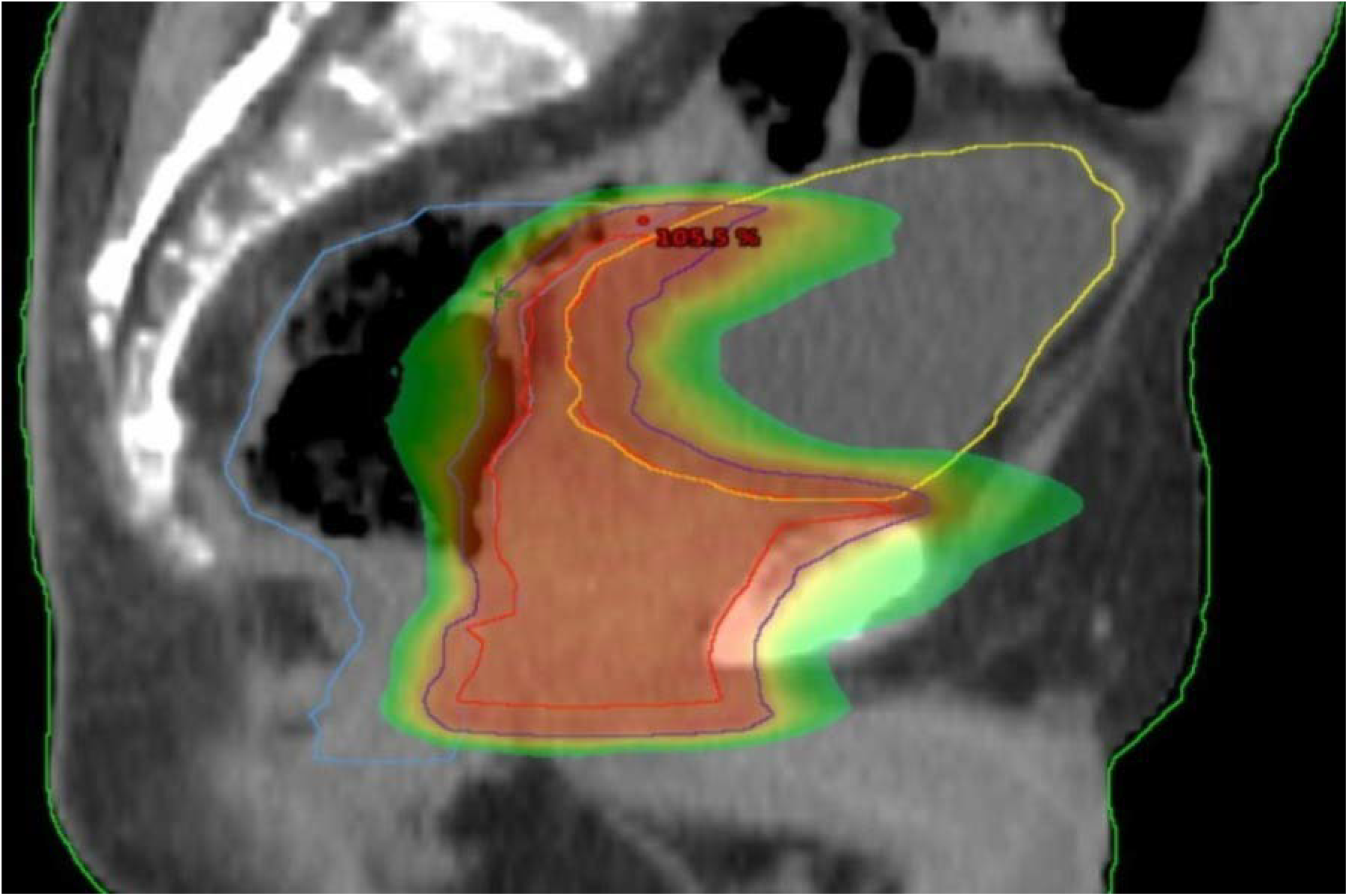
Pre-treatment sagittal CT image of the prostate showing 50% isodose line distribution.

**FIGURE 4:**
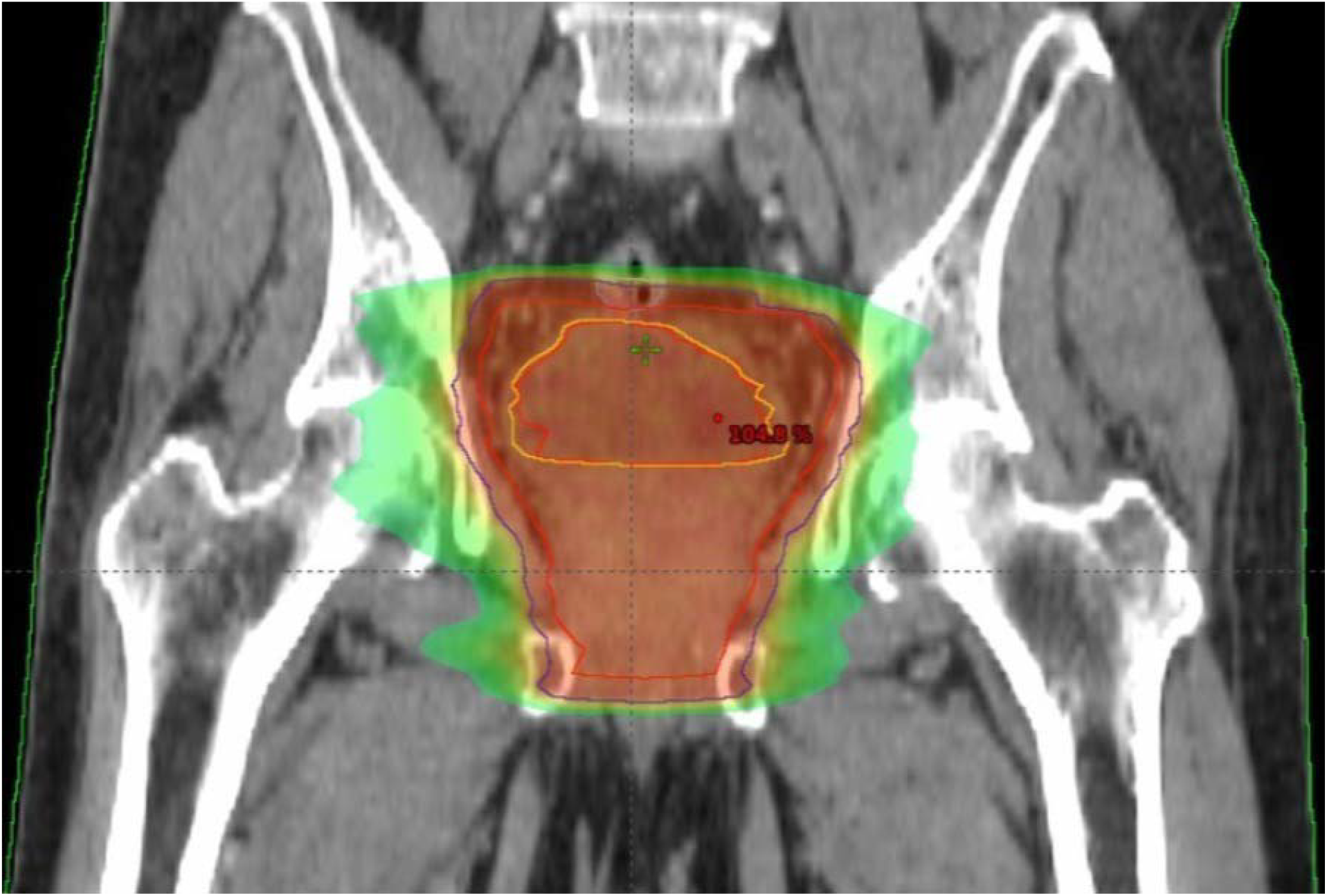
Pre-treatment coronal CT image of the prostate showing 50% isodose line distribution.

PTV D_95_ (35.80 Gy) is below prescription (36.25 Gy) with statistical significance (p<0.001) [9-14, 16-19]. But D_95_ covered 95% of the prescribed dose.Dmean/Dmax exceeds prescription (p<0.001), indicating hot spots(see Table 3). In SBRT, such hot spots are often intentionally created within the target to ensure adequate tumor coverage due to the highly conformal and hypofractionated nature of the treatment. These dose inhomogeneities can enhance tumor control by delivering a higher dose to the tumor core while maintaining normal tissue constraints. However, careful planning is essential to prevent excessive doses to surrounding tissues and minimize toxicity.

**TABLE 3:**
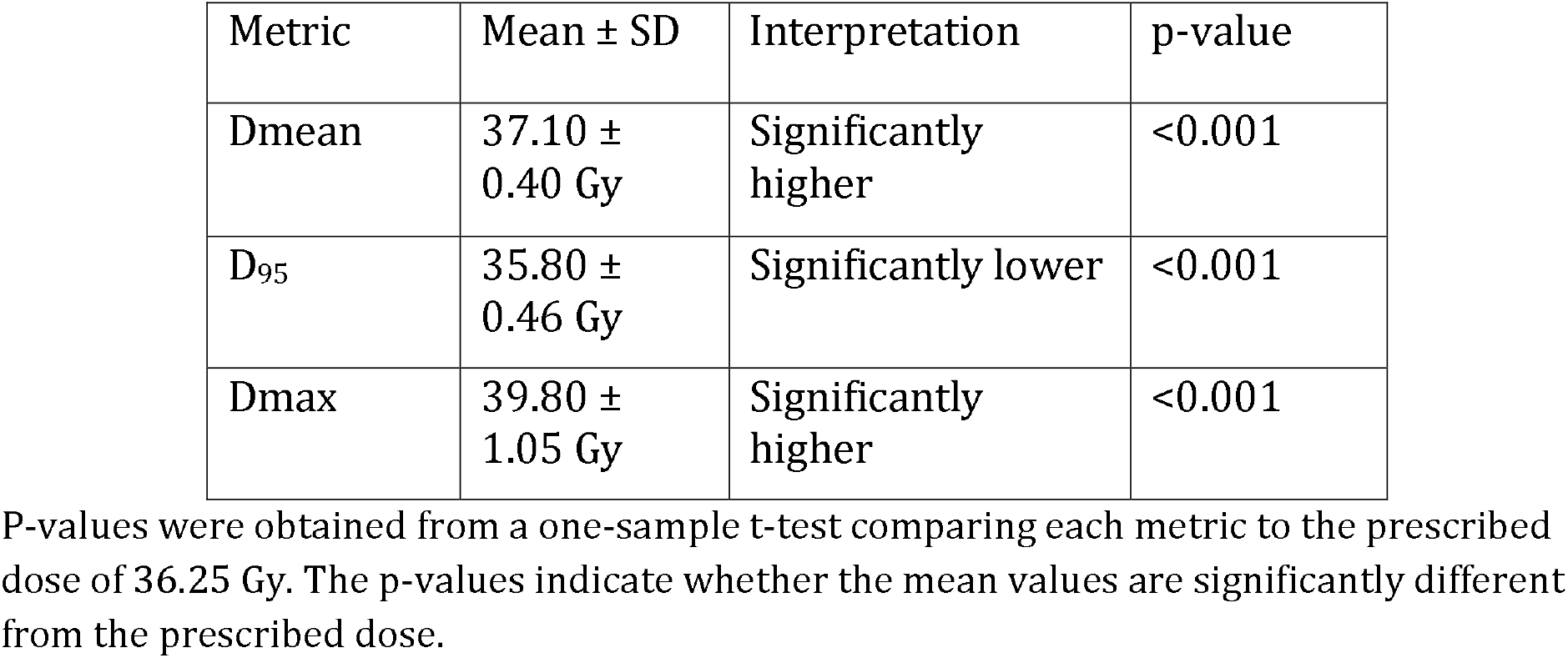
PTV Metrics vs Prescription Dose(36.25 Gy)

| Metric | Mean ± SD | Interpretation | p-value |
| --- | --- | --- | --- |
| Dmean | 37.10 ± 0.40 Gy | Significantly higher | <0.001 |
| D<sub>95</sub> | 35.80 ± 0.46 Gy | Significantly lower | <0.001 |
| Dmax | 39.80 ± 1.05 Gy | Significantly higher | <0.001 |

P-values were obtained from a one-sample t-test comparing each metric to the prescribed dose of 36.25 Gy. The p-values indicate whether the mean values are significantly different from the prescribed dose.

Table 4 illustrates the dosimetric analysis demonstrated high treatment accuracy, with mean gamma pass rates of 99.94±0.08% for 3%/2mm and 99.67±0.23% for 2%/2mm criteria. The conformity index and gradient index were 0.905±0.18 and 3.08±0.16, respectively, indicating precise target coverage and sharp dose fall-off.

**TABLE 4:** Gamma Analysis and Plan Quality Metrics for Prostate SBRT Plans.

| Metric | 3%/2 mm<br>Gamma Pass<br>Rate (%) | 2%/2 mm<br>Gamma Pass<br>Rate (%) | Conformity<br>Index (CI) | Gradient<br>Index (GI) |
| --- | --- | --- | --- | --- |
| Mean ±<br>SD | 99.94 ± 0.08 | 99.67 ± 0.23 | 0.905 ± 0.18 | 3.08 ± 0.16 |

Figure 5 offers visual insights into the balance between tumor coverage and sparing healthy organs during treatment planning. It includes four panels: (A) a box plot of PTV dose distribution; (B) the relationship between PTV V_95_ and bladder V_18Gy_; (C) PTV V_95_ versus penile bulb dose; and (D) PTV V_95_ versus rectum V_18Gy_. These panels collectively illustrate the various trade-offs involved in dose distribution.

**FIGURE 5:**
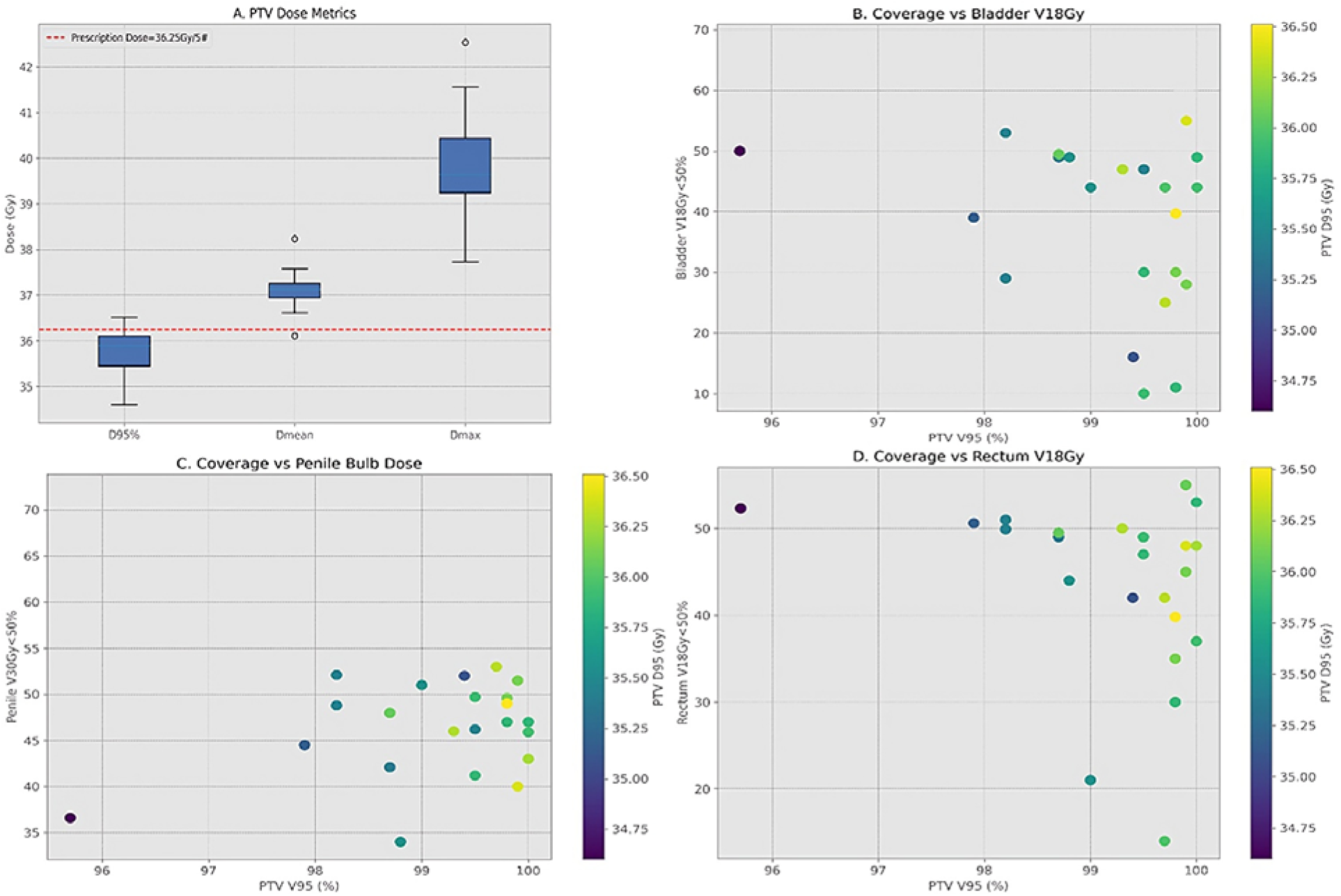
(A) Box plot of PTV dose distribution. (B) Relationship between PTV V_95_ and bladder V_18Gy_. (C) PTV V_95_ versus penile bulb dose. (D) PTV V_95_ versus rectum V_18Gy_.

**Figure 6:**
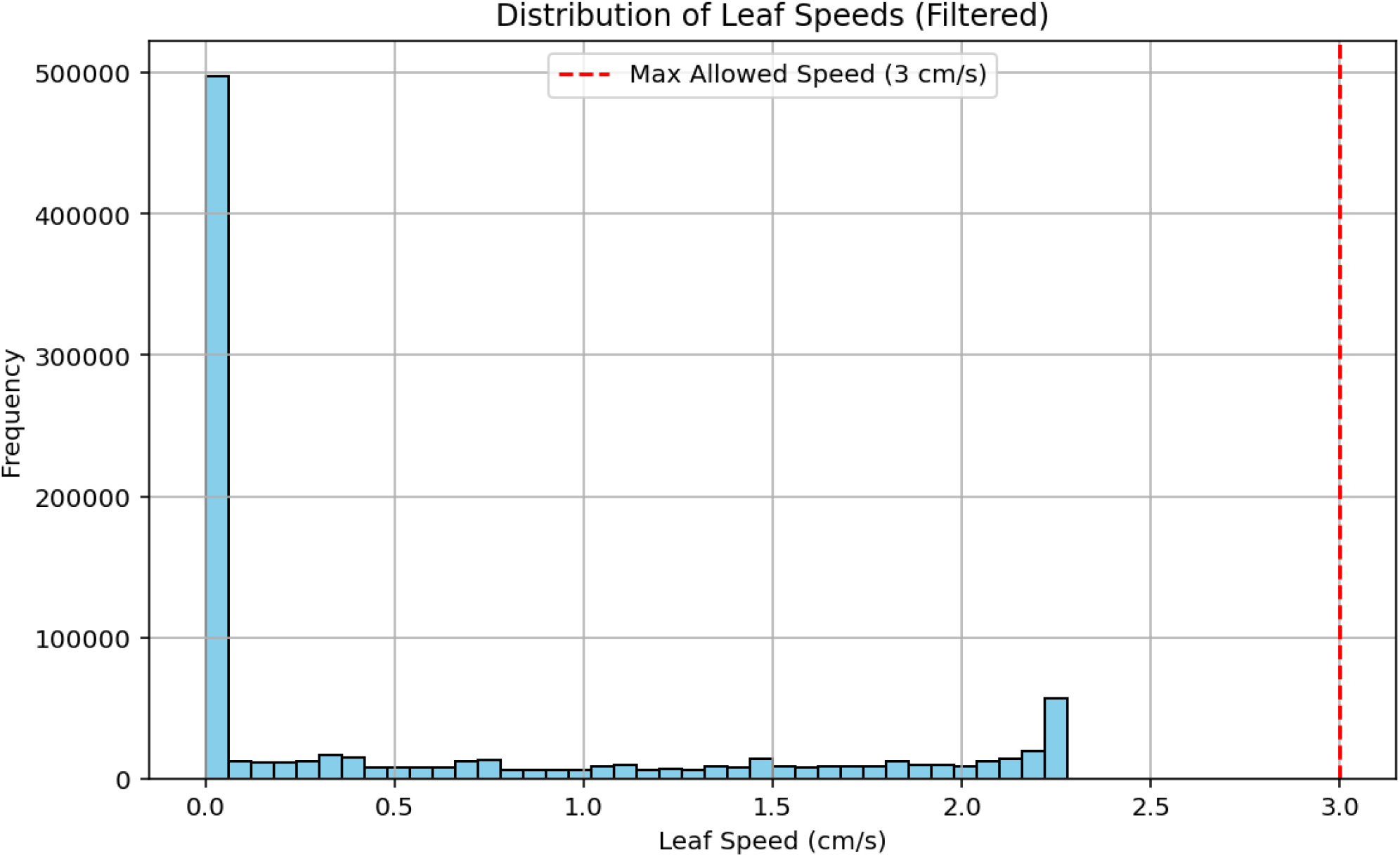
Distribution of MLC leaf speed data during treatment delivery, demonstrating variations in leaf velocity required for dose modulation and conformity.

**Figure 7:**
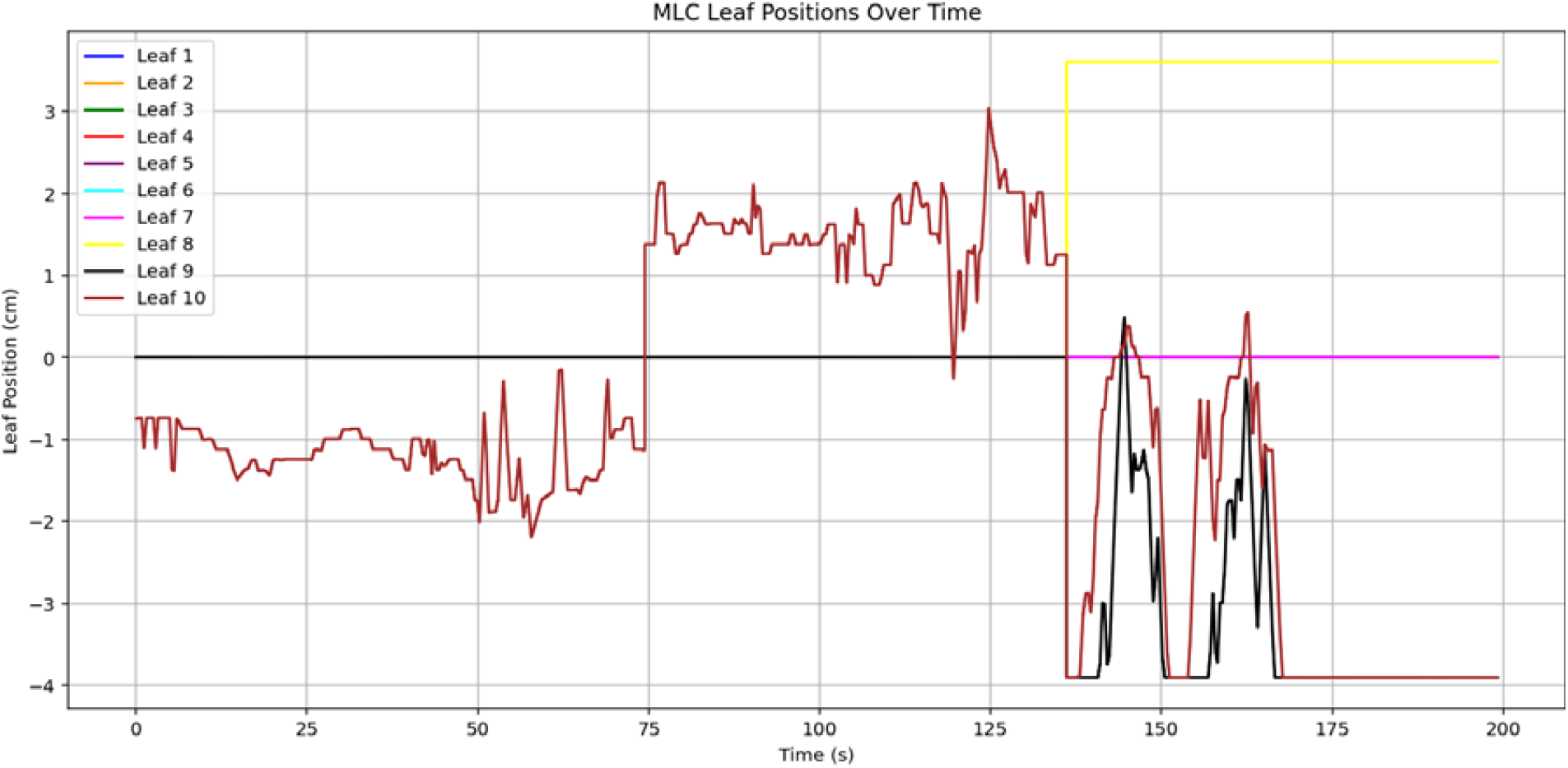
MLC leaf positions (leaves 1–10) over time during treatment, illustrating dynamic movements to shape the beam.

## 4. Discussion

### 4.1 Overview of Study

This study demonstrates that prostate SBRT delivered via RapidArc on a Varian Millennium MLC system achieves highly conformal and precise dose distributions, providing target coverage consistent with or exceeding established standards. The median PTV D_95%_ was 35.8 Gy, comparable to or slightly better than doses reported in recent clinical trials such as PACE-B, which documented an average of 34.7 ± 0.8 Gy. These results confirm that high-quality dose coverage is feasible with SBRT using RapidArc [3]. Organ at risk (OAR) parameters, including bladder Dmax (38.74 Gy), were marginally above some reported values like those from the PROFIT trial (38.5 ± 2.1 Gy). Nonetheless, these doses remained within acceptable toxicity thresholds, emphasizing that careful planning can balance tumor coverage with OAR sparing. The slight deviations highlight the importance of strict constraint application during planning and optimization.

### 4.2 Cohort Demographics and Their Implications

The relatively young mean age of our patient cohort (50 years) is an unusual finding, as prostate cancer is more commonly diagnosed in older men, with a median age of approximately 66 years. This age distribution may be influenced by referral patterns at our institution, where younger patients with intermediate-risk disease and good performance status are more frequently selected for SBRT. While younger patients may exhibit better tolerance to treatment-related side effects, their longer life expectancy underscores the importance of minimizing long-term toxicity and preserving quality of life post-SBRT.

### 4.3 Treatment Planning and Delivery Efficiency

The RapidArc approach enabled treatment times of approximately 70–90 seconds per arc, significantly reducing session duration compared to traditional methods. This rapid delivery minimizes the potential for intrafractional prostate motion; however, without real-time tracking systems, some residual uncertainties may persist. To mitigate intra-fraction motion, daily CBCT imaging was performed immediately before each arc to verify and, if necessary, correct patient positioning and prostate localization. While this protocol results in an increased imaging dose compared to single-CBCT protocols, the cumulative imaging dose was estimated to be less than **100–150**⍰**mGy per fraction**, which is considered acceptable relative to the total therapeutic dose of 36.25⍰Gy. Moreover, the improved geometric accuracy achieved through repeated imaging was deemed critical for high-precision SBRT.

Specifically, CBCT scans acquired before each arc allowed for precise alignment of the prostate relative to the planning CT. This intra-fraction imaging protocol facilitated real-time verification of treatment setup, ensuring accurate dose delivery during each arc. [9]. Although continuous intra-fraction motion tracking was not employed, this approach effectively minimized positional errors between arcs and maintained high dosimetric accuracy. Our dosimetric verification confirmed the consistent agreement between planned and delivered doses, underscoring the mechanical and dosimetric stability of the system and the effectiveness of the image-guided setup. The advanced inverse planning algorithms tailored for RapidArc enabled the generation of plans that met all clinical criteria, demonstrating the technique’s capacity for high-quality, conformal treatment delivery efficiently.

### 4.4 Comparison with Literature and Clinical Implications

Our dosimetric outcomes align well with findings from recent SBRT studies. For example, the PACE-B trial reported a prostate D_95%_ of approximately 34.7 Gy, similar to our results, confirming the adequacy of target coverage [1, 3]. The slightly higher bladder Dmax in our cohort suggests that further refinement of planning constraints could enhance OAR sparing. The combination of rapid treatment times and intra-fraction CBCT imaging before each arc effectively reduces setup uncertainties and potential prostate motion during treatment. This approach allows for immediate verification and correction, ensuring high precision in dose delivery without the need for continuous intra-fraction tracking devices [12]. Although some plans exhibited higher doses to the bladder, ongoing optimization of planning constraints—such as tighter OAR limits and adaptive planning—may further improve normal tissue sparing without compromising tumor coverage.

### 4.5 Limitations and Future Directions

This study’s limitations include its retrospective, single-institution design and a modest sample size (n=24). The absence of continuous intra-fraction motion tracking means that some prostate motion during treatment was not directly monitored, which could impact dose accuracy. Incorporating intra-fraction imaging or adaptive techniques, such as MR-guided radiotherapy, could further enhance treatment precision in future applications [12]. Exploring different arc configurations, dose escalation schemes, and adaptive planning approaches can optimize therapeutic outcomes. Larger, prospective studies are necessary to validate these findings and compare various motion management strategies.

#### Leaf Speed Distribution

As shown in Figure 6, the histogram of multileaf collimator (MLC) leaf speeds during treatment demonstrates that the majority of leaves remained stationary or moved very slowly, with speeds clustered near zero and a mean of 0.59⍰cm/s. Only a few instances of higher leaf speeds were observed, with a maximum of approximately 2.3⍰cm/s—well below the mechanical limit of 2.5⍰cm/s defined for the Varian Millennium 120 MLC system[21]. The narrow distribution suggests controlled and deliberate MLC motion, with minimal rapid transitions or complex modulation. This behavior reflects precise beam shaping and low risk of mechanical inaccuracies, thereby contributing to accurate and safe dose delivery [5]. Overall, the observed speed profile indicates that the plan was executed smoothly, consistent with a well-optimized and safety-focused treatment strategy.

### 4.6 Comparison of MLC Leaf Motion Relative to Isocenter (Center of PTV)

In this study, we performed an in-depth analysis of MLC leaf dynamics during radiotherapy delivery, focusing on the positional behavior of specific leaf groups relative to the isocenter, which was positioned at the center of the PTV (Planning Target Volume). We compared the movement patterns of the offset leaves (leaves 1 to 10) and the leaves located adjacent to the isocenter (leaves 30 to 40).

#### Offset Leaves (1-10)

As illustrated in Figure 7, these leaves, positioned away from the isocenter, showed relatively stable positions throughout the treatment duration. Leaf 8 and 9 remained nearly stationary, indicating minimal modulation, likely serving as fixed field boundaries. In contrast, leaf 10 demonstrated significant and rapid positional shifts after approximately 60 seconds, suggesting active modulation to accommodate treatment requirements or compensate for patient movement.

#### Leaves Adjacent to Isocenter (30-40)

Figure 8 depicts the movement patterns of the leaves located near the isocenter, which was placed at the center of the PTV. These leaves exhibited continuous fluctuations and abrupt positional changes over the course of the treatment session.This dynamic behavior reflects the necessity for precise modulation near the target to achieve conformal dose delivery while sparing surrounding healthy tissue.

**Figure 8:**
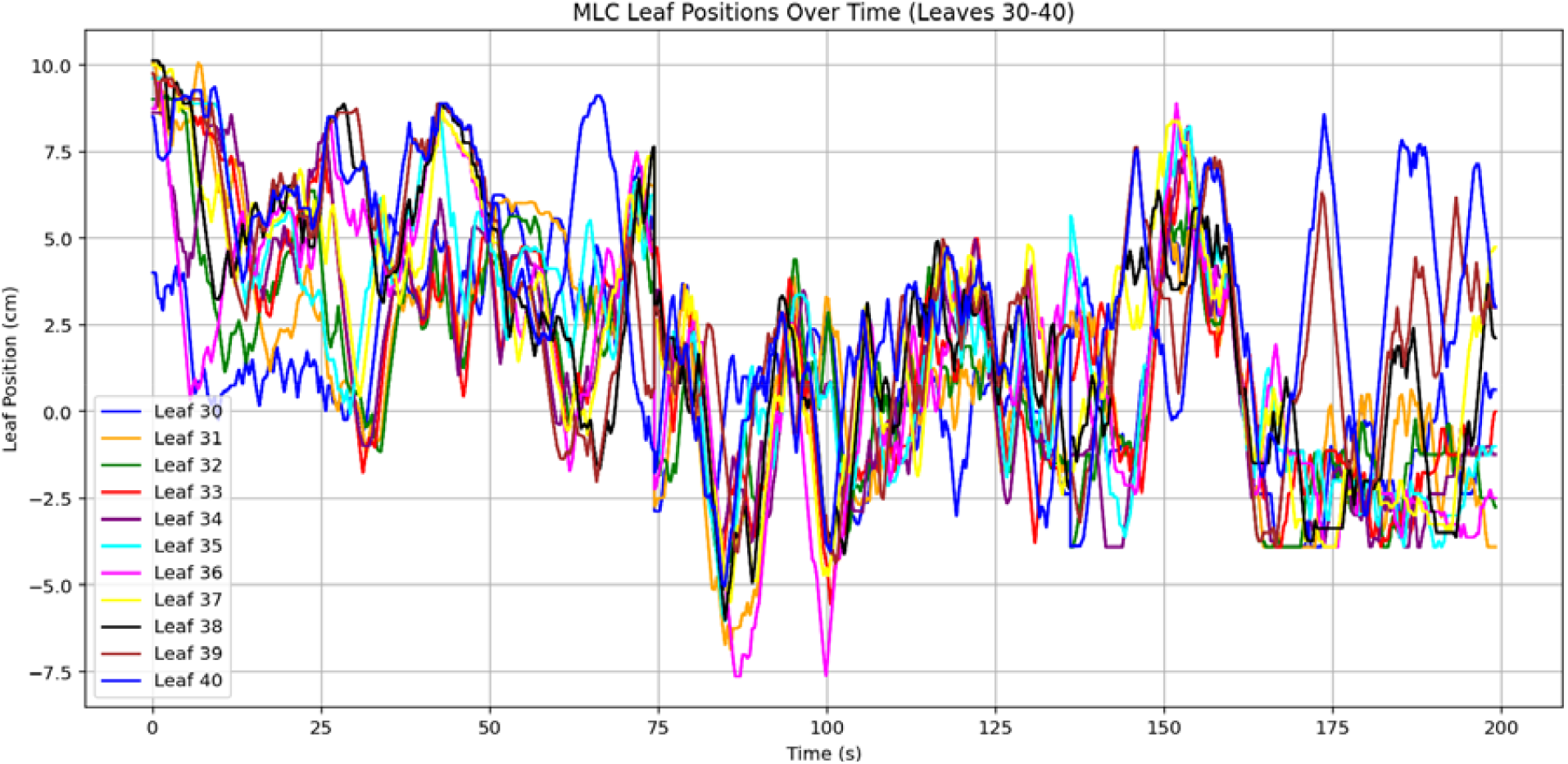
MLC leaf positions (leaves 30–40) over time, showing adjustments in outer bank leaves for beam shaping and dose control.

### 4.7 Study Strengths and Generalisability

This study is strengthened by its detailed dosimetric evaluation incorporating not only PTV and OAR metrics but also advanced parameters such as MLC leaf speed and dynamic leaf positioning, providing a comprehensive insight into treatment delivery accuracy. Additionally, the use of intra-fraction CBCT before each arc enhanced treatment precision without requiring continuous motion tracking, making the workflow efficient and clinically practical.

### 4.8 Clinical Significance

Our results support that SBRT delivered via RapidArc, with intra-fraction Cone Beam Computed Tomography(CBCT) imaging before each arc [9], offers a practical and effective treatment option for prostate cancer. This approach achieves high-quality conformal dose distributions with efficient treatment times and robust verification of prostate positioning during each arc, all without the need for real-time motion tracking devices. This protocol minimizes treatment duration and setup uncertainties, enhancing patient comfort and treatment accuracy. The intra-fraction CBCT verification before each arc ensures precise targeting and dose delivery, making this a reliable and accessible strategy in routine clinical practice. In conclusion, SBRT using RapidArc combined with intra-fraction CBCT imaging before each arc provides a promising approach for prostate radiotherapy, balancing precision, efficiency, and safety. Future advancements in intra-fraction motion management will further refine this modality and potentially improve clinical outcomes.

## 5. Conclusions

This study reinforces the value of SBRT using advanced delivery techniques such as RapidArc for the precise and efficient treatment of prostate cancer. Daily intra-fraction CBCT imaging before each arc enhances localization accuracy by effectively managing organ motion, ensuring optimal target coverage while minimizing dose to surrounding normal tissues. The implementation of high-dose, hypofractionated regimens—such as 36.25⍰Gy in five fractions—demonstrated excellent dosimetric feasibility and safety. A key contribution of this work is the detailed multileaf collimator (MLC) motion analysis, which revealed a predominantly slow and stable leaf speed distribution. The limited occurrence of high-speed MLC movements supports the mechanical accuracy and consistency of plan delivery. This added layer of verification complements conventional dosimetric metrics, providing further assurance of treatment quality and safety. While RapidArc significantly shortens treatment time and improves conformity, careful adherence to bladder and rectum dose constraints remains critical to reduce toxicity risks. These findings support the continued advancement of personalized, high-precision radiotherapy strategies. Future studies should further refine motion management protocols and incorporate real-time mechanical metrics, such as MLC behavior, to enhance outcome predictability in prostate SBRT.

## Data Availability

All data produced in the present study are available upon reasonable request to the authors

